# Assessment of N95 respirator decontamination and re-use for SARS-CoV-2

**DOI:** 10.1101/2020.04.11.20062018

**Authors:** Robert J. Fischer, Dylan H. Morris, Neeltje van Doremalen, Shanda Sarchette, M. Jeremiah Matson, Trenton Bushmaker, Claude Kwe Yinda, Stephanie N. Seifert, Amandine Gamble, Brandi N. Williamson, Seth D. Judson, Emmie de Wit, James O. Lloyd-Smith, Vincent J. Munster

## Abstract

The unprecedented pandemic of SARS-CoV-2 has created worldwide shortages of personal protective equipment, in particular respiratory protection such as N95 respirators. SARS-CoV-2 transmission is frequently occurring in hospital settings, with numerous reported cases of nosocomial transmission highlighting the vulnerability of healthcare workers. In general, N95 respirators are designed for single use prior to disposal. Here, we have analyzed four readily available and often used decontamination methods: UV, 70% ethanol, 70C heat and vaporized hydrogen peroxide for inactivation of SARS-CoV-2 on N95 respirators. Equally important we assessed the function of the N95 respirators after multiple wear and decontamination sessions.

---

Dear editor,

The unprecedented pandemic of COVID-19 has created worldwide shortages of personal protective equipment, in particular respiratory protection such as N95 respirators(1). SARS-CoV-2 transmission is frequently occurring in hospital settings, with numerous reported cases of nosocomial transmission highlighting the vulnerability of healthcare workers(2). The environmental stability of SARS-CoV-2 underscores the need for rapid and effective decontamination methods. In general, N95 respirators are designed for single use prior to disposal. Extensive literature is available for decontamination procedures for N95 respirators, using either bacterial spore inactivation tests, bacteria or respiratory viruses (e.g. influenza A virus)(3-6). Effective inactivation methods for these pathogens and surrogates include UV, ethylene oxide, vaporized hydrogen peroxide (VHP), gamma irradiation, ozone and dry heat(3-7). The filtration efficiency and N95 respirator fit has typically been less well explored, but suggest that both filtration efficiency and N95 respirator fit can be affected by the decontamination method used(7, 8). For a complete list of references see supplemental information.

Here, we analyzed four different decontamination methods – UV radiation (260 – 285 nm), 70°C dry heat, 70% ethanol and vaporized hydrogen peroxide (VHP) – for their ability to reduce contamination with infectious SARS-CoV-2 and their effect on N95 respirator function. For each of the decontamination methods, we compared the normal inactivation rate of SARS-CoV-2 on N95 filter fabric to that on stainless steel, and we used quantitative fit testing to measure the filtration performance of the N95 respirators after each decontamination run and 2 hours of wear, for three consecutive decontamination and wear sessions (see supplemental information). VHP and ethanol yielded extremely rapid inactivation both on N95 and on stainless steel (Figure 1A). UV inactivated SARS-CoV-2 rapidly from steel but more slowly on N95 fabric, likely due its porous nature. Heat caused more rapid inactivation on N95 than on steel; inactivation rates on N95 were comparable to UV.

**Figure 1.**
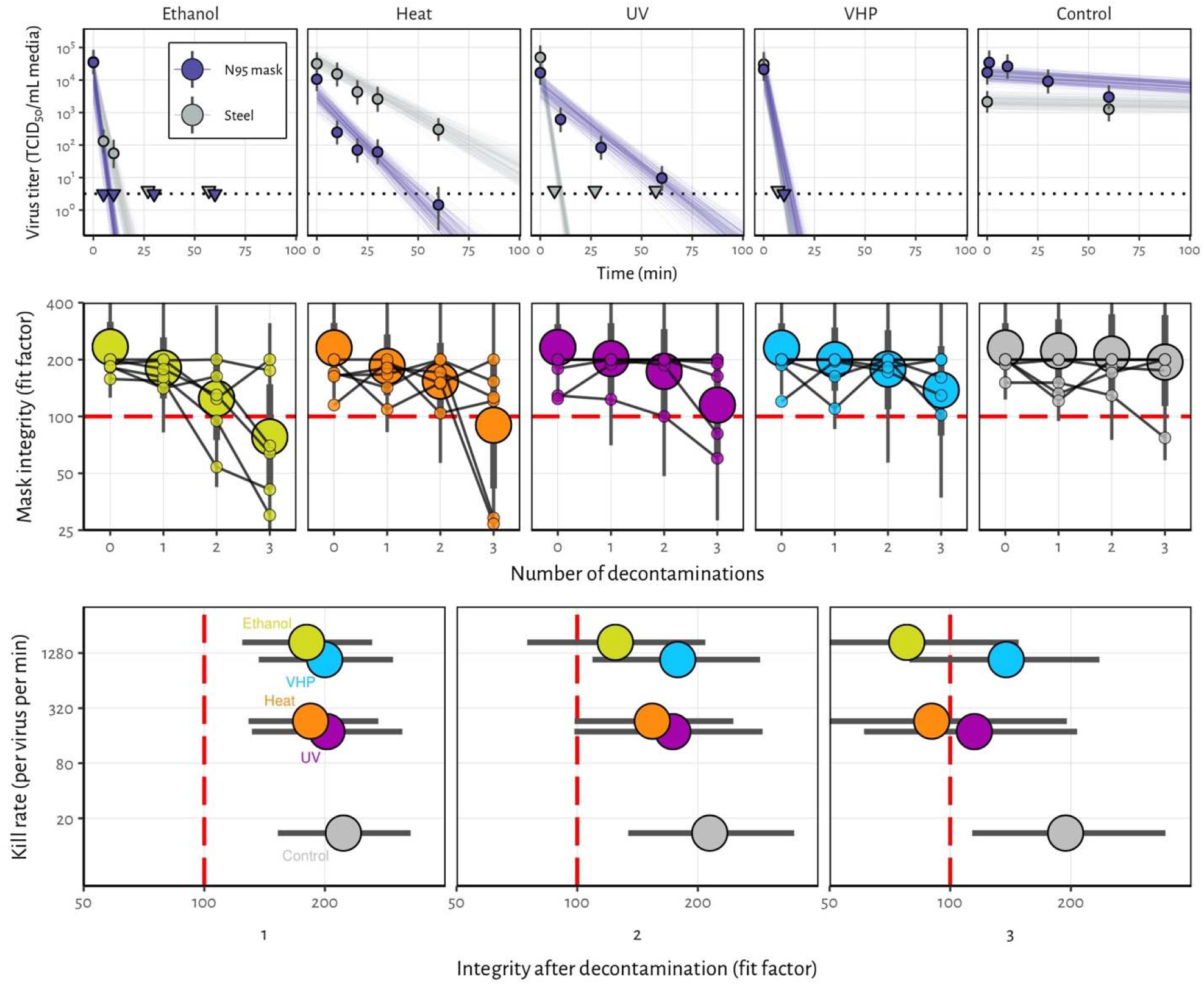
Decontamination of SARS-CoV-2 by four different methods. **A)** SARS-CoV-2 on N95 fabric and stainless steel surface was exposed to UV, 70 °C dry heat, 70% ethanol and vaporized hydrogen peroxide (VHP). 50 µl of 10^5^ TCID_50_/mL of SARS-CoV was applied on N95 and stainless steel (SS). Samples were collected at indicted time-points post exposure to the decontamination method for UV, heat and ethanol and after 10 minutes for VHP. Viable virus titer is shown in TCID_50_/mL media on a logarithmic scale. All samples were quantified by end-point titration on Vero E6 cells. Plots show estimated mean titer across three replicates (circles and bars show the posterior median estimate of this mean and a 95% credible interval). Time-points with no positive wells for any replicate are plotted as triangles at the approximate single-replicate detection limit of the assay (LOD, see Appendix for discussion) to indicate that a range of sub-LOD values are plausible. Steel points at the LOD are offset slightly up and to the left to avoid overplotting. Lines show predicted decay of virus titer over time (lines; 50 random draws per replicate from the joint posterior distribution of the exponential decay rate, i.e. negative of the slope, and intercept, i.e. initial virus titer). Black dotted line shows approximate LOD: 10_0.5_ TCID_50_/mL media. **B)** Mask integrity. Quantitative fit testing results for all the decontamination methods after decontamination and 2 hours of wear, for three consecutive runs. Data from six individual replicates (small dots) for each treatment are shown in addition to an estimated median fit factor (large dots), an estimated 68% range of fit factors (thick bars) and an estimated 95% range (thin bars). Fit factors are a measure of filtration performance: the ratio of the concentration of particles outside the mask to the concentration inside. The measurement machine reports value up to 200. A minimal fit factor of 100 (red dashed line) is required for a mask to pass a fit test. **C)** SARS-CoV-2 decontamination performance. Kill rate (y-axis), versus mask integrity after decontamination (x-axis; point represents estimated median, bar length represents estimated 68% range). The three panels report mask integrity after one, two or three decontamination cycles.

Quantitative fit tests showed that the filtration performance of the N95 respirator was not markedly reduced after a single decontamination for any of the four decontamination methods (Figure 1B). Subsequent rounds of decontamination caused sharp drops in filtration performance of the ethanol-treated masks, and to a slightly lesser degree, the heat-treated masks. The VHP and UV treated masks retained comparable filtration performance to the control group after two rounds of decontamination, and maintained acceptable performance after three rounds.

Taken together, our findings show that VHP treatment exhibits the best combination of rapid inactivation of SARS-CoV-2 and preservation of N95 respirator integrity, under the experimental conditions used here (Figure 1C). UV radiation kills the virus more slowly and preserves comparable respirator function. 70°C dry heat kills with similar speed to UV and is likely to maintain acceptable fit scores for two rounds of decontamination. Ethanol decontamination is not recommended due to loss of N95 integrity, echoing earlier findings^5^.

All treatments, particularly UV and dry heat, should be conducted for long enough to ensure that a sufficient reduction in virus concentration has been achieved. The degree of required reduction will depend upon the degree of initial virus contamination. Policymakers can use our estimated decay rates together with estimates of real-world contamination to choose appropriate treatment durations (see supplemental information).

Our results indicate that N95 respirators can be decontaminated and re-used in times of shortage for up to three times for UV and HPV, and up to two times for dry heat. However, utmost care should be given to ensure the proper functioning of the N95 respirator after each decontamination using readily available qualitative fit testing tools and to ensure that treatments are carried out for sufficient time to achieve desired risk-reduction. It will therefore be critical that FDA, CDC and OSHA guidelines with regards to fit testing, seal check and respirator re-use are followed(9, 10).

## Data Availability

Code and data to reproduce the Bayesian estimation results and produce corresponding figures are archived online at OSF: and available on Github:

## Supplemental methods

### Short literature review on mask decontamination

The COVID-19 pandemic has highlighted the necessity for large-scale decontamination procedures for personal protective equipment (PPE), in particular N95 respirator masks(1). SARS-CoV-2 has frequently been detected on PPE of healthcare workers(11). The environmental stability of SARS-CoV-2 underscores the need for rapid and effective decontamination methods(12). Extensive literature is available for decontamination procedures for N95 respirators, using either bacterial spore inactivation tests, bacteria or respiratory viruses (e.g. influenza A virus)(3-6, 9, 13-15). Effective inactivation methods for these pathogens and surrogates include UV, ethylene oxide, vaporized hydrogen peroxide (VHP), gamma irradiation, ozone and dry heat(4, 5, 7, 9, 14-16). The filtration efficiency and N95 respirator fit has typically been less well explored, but suggest that both filtration efficiency and N95 respirator fit can be affected by the decontamination method used(7, 8). It will therefore be critical that FDA, CDC and OSHA guidelines with regards to fit testing, seal check and respirator re-use are followed(9, 17-20).

### Laboratory experiments

#### Viruses and titration

HCoV-19 nCoV-WA1-2020 (MN985325.1) was the SARS-CoV-2 strain used in our comparison(21). Virus was quantified by end-point titration on Vero E6 cells as described previously(22). Virus titrations were performed by end-point titration in Vero E6 cells. Cells were inoculated with 10-fold serial dilutions in four-fold of samples taken from N95 mask and stainless steel surfaces (see below). One hour after inoculation of cells, the inoculum was removed and replaced with 100 µl (virus titration) DMEM (Sigma-Aldrich) supplemented with 2% fetal bovine serum, 1 mM L-glutamine, 50 U/ml penicillin and 50 µg/ml streptomycin. Six days after inoculation, cytopathogenic effect was scored and the TCID_50_ was calculated (see below). Wells presenting cytopathogenic effects due to media toxicity (e.g., due to the presence of ethanol or hydrogen peroxide) rather than viral infection were removed from the titer inference procedure.

#### N95 and stainless steel surface

N95 material discs were made by punching 9/16” (15 mm) fabric discs from N95 respirators, AOSafety N9504C respirators (Aearo Company Southbridge, MA). The stainless steel 304 alloy discs were purchased from Metal Remnants (https://metalremnants.com/) as described previously. 50 µL of SARS-CoV-2 was spotted onto each disc. A 0 time-point measurement was taken prior to exposing the discs to the disinfection treatment. At each sampling time-point, discs were rinsed 5 times by passing the medium over the stainless steel or through the N95 disc. The medium was transferred to a vial and frozen at −80°C until titration. All experimental conditions were performed in triplicate.

#### Decontamination methods

##### Ultraviolet light

Plates with fabric and steel discs were placed under an LED high power UV germicidal lamp (effective UV wavelength 260-285nm) without the titanium mesh plate (LEDi2, Houston, Tx) 50 cm from the UV source. At 50 cm the UVC power was measured by the manufacturer at 550 µW/cm^2^. Plates were removed at 10, 30 and 60 minutes and 1 mL of cell culture medium added. The energy the discs were exposed to at 10, 30 and 60 min is 0.33 J/cm^2^, 0.99 J/cm^2^, and 1.98 J/cm^2^ respectively. While the CDC has no specific recommendations on the minimum dose, they do report that a 1 J/cm2 dose can reduce tested viable viral loads by 99.9%^4^.

##### Heat treatment

Plates with fabric and steel discs were placed in a 70°C oven. Plates were removed at 10, 20, 30 and 60 minutes and 1 mL of cell culture medium added.

##### 70% ethanol

Fabric and steel discs were placed into the wells of one 24 well plate per time-point and sprayed with 70% ethanol to saturation. The plate was tipped to near vertical and 5 passes of ethanol were sprayed onto the discs from approximately 10 cm. After 10 minutes, 1 mL of cell culture medium was added.

##### VHP

Plates with fabric and steel discs were placed into a Panasonic MCO-19AIC-PT (PHC Corp. of North America Wood Dale, IL) incubator with VHP generation capabilities and exposed to hydrogen peroxide (approximately 1000 ppm). The exposure to VHP was 7 minutes, after the inactivation of the hydrogen peroxide, the plate was removed and 1 mL of cell culture medium was added.

##### Control

Plates with fabric and steel discs and steel plates were maintained at 21-23°C and 40% relative humidity for up to four days. After the designated time-points, 1 mL of cell culture medium was added.

#### N95 mask integrity testing

N95 Mask (3M™ Aura™ Particulate Respirator 9211+/37193) integrity testing after 2 hours of wear and decontamination, for three consecutive rounds, was performed for a total of 6 times for each decontamination condition and control condition. Masks were worn by subjects and integrity was quantitatively determined using the Portacount Respirator fit tester (TSI, 8038) with the N95 companion component, following the modified ambient aerosol condensation nuclei counter quantitative fit test protocol approved by the OSHA^18^. Subjects were asked to bend over for 40 seconds, talk for 50 seconds, move head from side-to-side for 50 seconds, and move head up-and-down for 50 seconds whilst aerosols on inside and outside of mask were measured. By convention, this fit test is passed when the final score is ≥100. For the N95 integrity testing, a Honeywell Mistmate humidifier (cat#HUL520B) was used for particle generation.

### Statistical analyses

In the model notation that follows, the symbol ∼ denotes that a random variable is distributed according to the given distribution. Normal distributions are parametrized as Normal(mean, standard deviation). Positive-constrained normal distributions (“Half-Normal”) are parametrized as Half-Normal(mode, standard deviation). Normal distributions truncated to the interval [0, 1] are parameterized as TruncNormal(mode, standard deviation).

We use <Distribution Name>CDF(x | parameters) and <Distribution Name>CCDF to denote the cumulative distribution function and complementary cumulative distribution functions of a probability distribution, respectively. So for example NormalCDF(5 | 0, 1) is the value of the Normal(0, 1) cumulative distribution function at 5.

We use logit(*x*) and invlogit(*x*) to denote the logit and inverse logit functions, respectively:

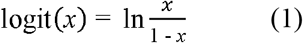

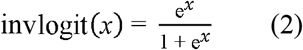

#### Mean titer inference

We inferred mean titers across sets of replicates using a Bayesian model. The log_10_ titers *v*_*ijk*_ (the titer for the sample from replicate *k* of time-point *j* of experiment *i*) were assumed to be normally distributed about a mean *µ*_*ij*_ with a standard deviation *σ*. We placed a very weakly informative normal prior on the log_10_ mean titers *µ*_*ij*_:

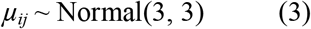

We placed a weakly informative normal prior on the standard deviation:

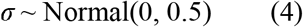

We then modeled individual positive and negative wells for sample *ijk* according to a Poisson single-hit model(23). That is, the number of virions that successfully infect cells in a given well is Poisson distributed with mean:

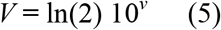

where *v* is the log_10_ virus titer in TCID_50_, where *v* is the log_10_ virus titer in TCID_50_, and the well is infected if at least one virion successfully infects a cell. The value of the mean derives from the fact that our units are TCID_50_; the probability of infection at *v* = 0, i.e. 1 TCID_50_, is equal to 1 – e ^-ln(2) × 1^ = 0.5.

Let *Y*_*ijkdl*_ be a binary variable indicating whether the *l*^th^ well of dilution factor *d* (expressed as log_10_ dilution factor) of sample *ijk* was positive (so *Y*_*ijkdl*_ = 1 if the well was positive and 0 otherwise), which will occur as long as at least one virion successfully infects a cell.

It follows from (5) that the conditional probability of observing *Y*_ijkdl_ = 1 given a true underlying titer log_10_ titer *v*_*ijk*_ is given by:

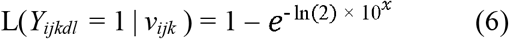

Where

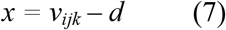

is the expected concentration, measured in log_10_ TCID_50_, in the dilute sample. This is simply the probability that a Poisson random variable with mean (– ln(2) × 10^*x*^) is greater than 0. Similarly, the conditional probability of observing *Y*_*ijkdl*_ = 0 given a true underlying titer log_10_ titer *v*_*ijk*_ is given by:

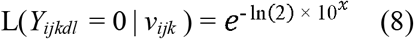

which is the probability that the Poisson random variable is 0.

This gives us our likelihood function, assuming independence of outcomes across wells.

#### Virus inactivation regression

The durations of detectability depend on the decontamination treatment but also initial inoculum and sampling method, as expected. We therefore estimated the decay rates of viable virus titers using a Bayesian regression analogous to that used in van Doremalen et al., 2020(12). This modeling approach allowed us to account for differences in initial inoculum levels across replicates as well as other sources of experimental noise. The model yields estimates of posterior distributions of viral decay rates and half-lives in the various experimental conditions – that is, estimates of the range of plausible values for these parameters given our data, with an estimate of the overall uncertainty(24).

Our data consist of 10 experimental conditions: 2 materials (N95 masks and stainless steel) by 5 treatments (no treatment, ethanol, heat, UV and VHP). Each has three replicates, and multiple time-points for each replicate. We analyze the two materials separately. For each, we denote by *Y*_*ijkdl*_ the positive or negative status (see above) for well *l* which has dilution *d* for the titer *v*_*ijk*_ from experimental condition *i* during replicate *j* at time-point *k*.

We model each replicate *j* for experimental condition *i* as starting with some true initial log_10_ titer *v*_*ij*_(0) = *v*_*ij*0_. We assume that viruses in experimental condition *i* decay exponentially at a rate λ_*i*_ over time *t*. It follows that:

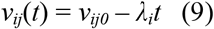

We use the direct-from-well data likelihood function described above, except that now instead of estimating titer distribution about a shared mean *µ*_*ij*_ we estimate λ_*i*_ under the assumptions that our observed well data *Y*_*ijkdl*_ reflect the titers *v*_*ij*_(*t*).

#### Regression prior distributions

We place a weakly informative Normal prior distribution on the initial log_10_ titers *v*_*ij0*_ to rule out implausibly large or small values (e.g. in this case undetectable log_10_ titers or log_10_ titers much higher than the deposited concentration), while allowing the data to determine estimates within plausible ranges:

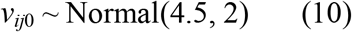

We placed a weakly informative Half-Normal prior on the exponential decay rates λ_*i*_:

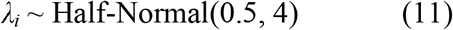

Our plated samples were of volume 0.1 mL, so inferred titers were incremented by 1 to convert to units of log_10_ TCID_50_/mL.

#### Mask integrity estimation

To quantify the decay of mask integrity after repeated decontamination, we used a logit-linear spline Bayesian regression to estimate the rate of degradation of mask fit factors over time, accounting for the fact that fit factors are interval-censored ratios. Fit factors are defined as the ratio of exterior concentration to interior concentration of a test aerosol. They are reported to the nearest integer, up to a maximum readout of 200, but arbitrarily large true fit factors are possible as the mask performance approaches perfect filtration.

We had 6 replicate masks *j* for each of 5 treatments *i* (no decontamination, ethanol, heat, UV and VHP). Each mask *j* was assessed for fit factor at 4 time-points *k*: before decontamination, and then after 1, 2, and 3 decontamination cycles. We label the control treatment *i =* 0. So we denote by *F*_*ijk*_ the fit factor for the *j*^th^ mask from the *i*^th^ treatment after *k* decontaminations (with *k =* 0 for the initial value).

We first converted fit factors *F*_*ijk*_ to the equivalent observed filtration rate *Y*_*ijk*_ by:

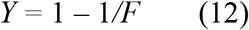

### Observation model and likelihood function

We modeled the censored observation process as follows. logit(*Y*_*ijk*_) values are observed with Gaussian error about the true filtration logit(*p*_*ijk*_), with an unknown standard deviation *σ*_*o*_, and then converted to fit factors, which are then censored:

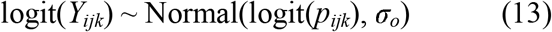

Because our reported fit factors are known to be within integer values and right-censored at 200, for *F*_*ijk*_ ≥ 200 we have a conditional probability of observing the data given the parameters of

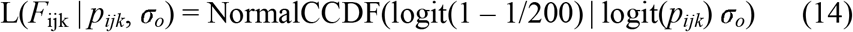

That is, we calculate the probability of observing a value of *F* greater than or equal to 200 (equivalent a value of *Y* greater than or equal to 1 – 1/200), given our parameters.

For 1.5 ≤ *F*_*ijk*_ *<* 200, we first calculate the upper and lower bounds of our observation *Y*^*+*^_*ijk*_ = 1 – 1 / (*F*_ijk_ – 0.5) and *Y*^*–*^_*ijk*_ = 1 – 1 / (*F*_*ijk*_ *–* 0.5). Then:

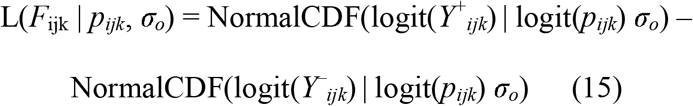

That is, we calculate the probability of observing a value between *Y*^*+*^_*ijk*_ and *Y*^*–*^_*ijk*_, given our parameters.

### Decay model

We assumed that each mask had some true initial filtration rate *p*_*ij*0_. We assumed that these were logit-normally distributed about some unknown mean mask initial filtration rate *p*_*avg*_ with a standard deviation *σ*_*p*_, that is:

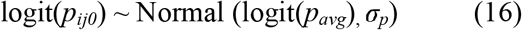

We then assumed that the logit of the filtration rate, logit(*p*_*ijk*_), decreased after each decontamination by a quantity *d*_0*k*_ *+ d*_*ik*_, where *d*_0*k*_ is natural degradation during the *k*^th^ trial in the absence of decontamination (i.e. the degradation rate in the control treatment, *i =* 0), and *d*_*ik*_ is the additional degrading effect of the *k*^th^ decontamination treatment of type *i >* 0). So for *k* = 1, 2, 3 and *i >* 0:

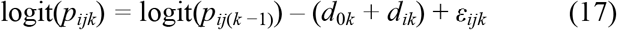

where *ε*_*ijk*_ is a normally-distributed error term with an inferred standard deviation *σ*_*εik*_ for each treatment and decontamination level.

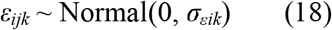

And for the control *i =* 0:

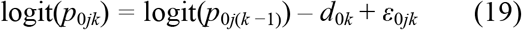

### Model prior distributions

We placed a weakly informative Half-Normal prior on the control degradation rate *d*_*0*_:

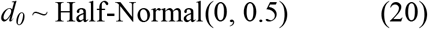

We placed a weakly informative Half-Normal prior on the non-control degradation rates *d*_*i*_, *i >* 0:

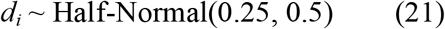

reflecting the conservative assumption that decontamination should degrade the mask at least somewhat.

We placed a Truncated Normal prior on the mean initial filtration *p*_*avg*_:

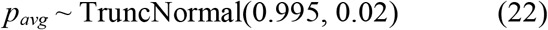

The mode of 0.995 corresponds to the maximum measurable fit factor of 200. The standard deviation of 0.02 leaves it plausible that some masks could start near or below the minimum acceptable threshold fit factor of 100, which corresponds to a *p* of 0.99.

We placed weakly informative Half-Normal priors on the logit-space standard deviations *σ*_*p*_, *σ*_*εik*_, and *σ*_*o*_. *σ*_*p*_ reflects variation in individual masks’ initial filtration about *p*_*avg*_. The various *σ*_*εik*_ reflect variation in masks’ true degree of degradation between decontaminations about the expected degree of decay, and *σ*_*o*_ reflects noise in the observation process.

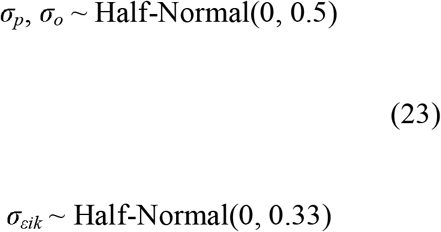

We chose standard deviations less than or equal to 0.5 for these normal hyperpriors because a standard deviation of 1.5 (i.e. 3 *σ* in the hyperprior) in logit space corresponds to probability values being uniformly distributed between 0 and 1; we therefore wish to tell our model not to use larger values of *σ*_*p*_, *σ*_*o*_, or *σ*_*εik*_, as these would squash all *p*_*ijk*_ to one of two modes, one at 0 and one at 1(25).

#### Markov Chain Monte Carlo Methods

For all Bayesian models, we drew posterior samples using Stan (Stan Core Team 2018), which implements a No-U-Turn Sampler (a form of Markov Chain Monte Carlo), via its R interface RStan. We ran four replicate chains from random initial conditions for 2000 iterations, with the first 1000 iterations as a warmup/adaptation period. We saved the final 1000 iterations from each chain, giving us a total of 4000 posterior samples. We assessed convergence by inspecting trace plots and examining _RD_ and effective sample size (*n*_*eff*_) statistics.

#### Limit of detection (LOD)

End-point titration has an approximate limit of detection set by the volume of the undilute sample deposited in each well. If all wells – including those containing undiluted sample – are negative and a Poisson single-hit model is used, the best guess is that the true titer lies somewhere below 1 TCID_50_ / (volume of deposited sample). How far below is determined by the number of wells. For four wells, as was standard in our experiments, the first quarter log_10_ titer at which 0 wells is the most likely outcome is 10^−0.5^ TCID_50_ per volume of sample. This is also the imputed Speaman-Karber titer in that case. Since we used samples of volume 0.1 mL, this corresponds to a value of 10^0.5^ TCID_50_/mL. So although we do not use the Spearman-Karber method here (since we infer mean titers directly from the well data) we use that LOD value to plot samples for which no replicate had a positive well (since the posterior distribution in that case covers a wide-range of sub-threshold values).

## Supplemental table

**Table S1.**
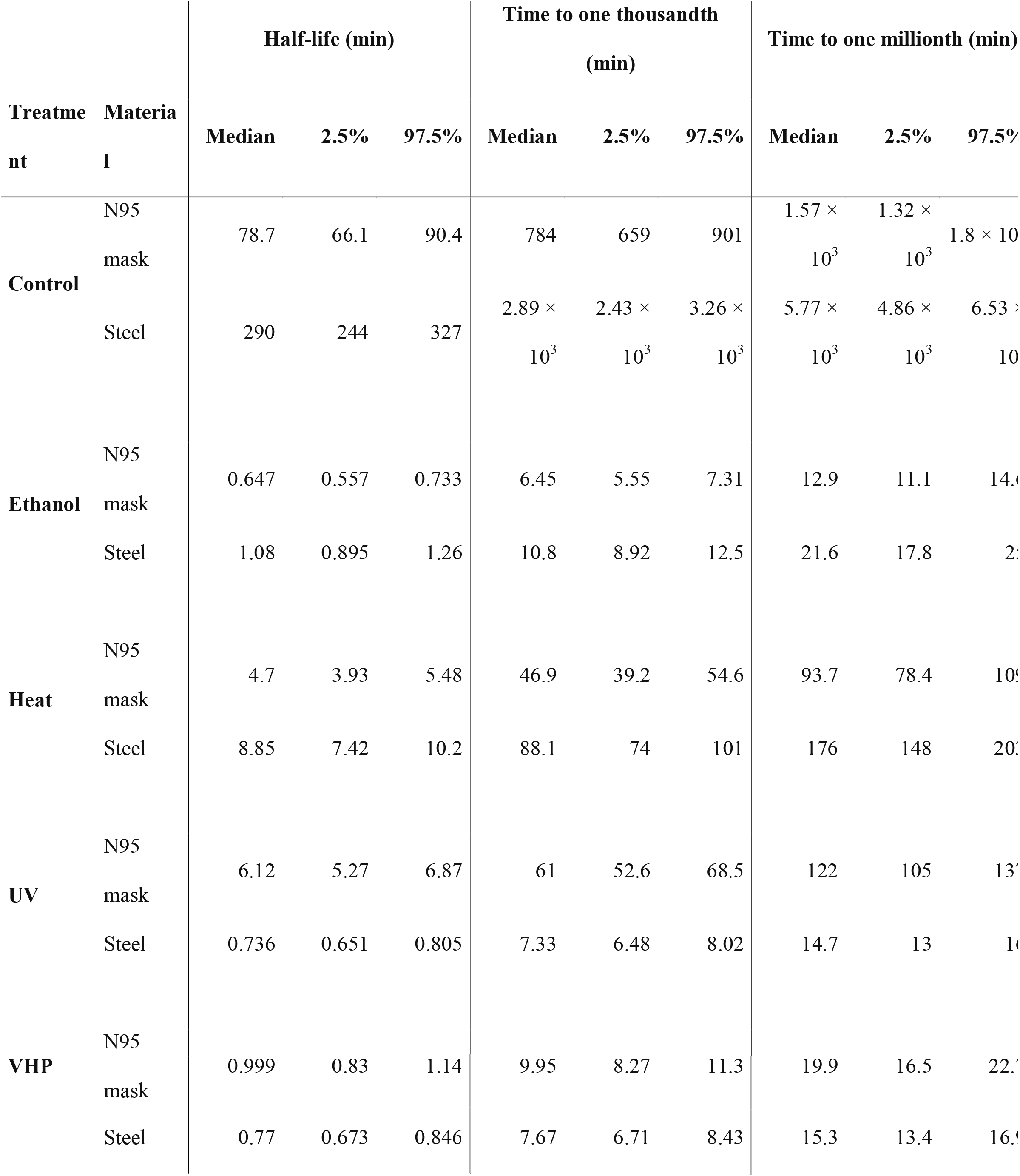
Effect of decontamination method on SARS-CoV-2 viability. Results are reported as the median and upper- and lower-limits of the 95% credible interval of the estimated half-life, and time needed to reduce viable SARS-CoV-2 load by a factor of one thousand or one million, based on the posterior distribution of the exponential decay rate of the virus on different materials following different decontamination treatments.

## Code and data availability

Code and data to reproduce the Bayesian estimation results and produce corresponding figures are archived online at OSF: https://doi.org/10.17605/OSF.IO/mkg9b and available on Github: https://github.com/dylanhmorris/n95-decontamination

## Acknowledgements

We would like to thank Madison Hebner, Julia Port, Kimberly Meade-White, Irene Offei Owusu, Victoria Avanzato and Lizzette Perez-Perez for excellent technical assistance. This research was supported by the Intramural Research Program of the National Institute of Allergy and Infectious Diseases (NIAID), National Institutes of Health (NIH). JOL-S and AG were supported by the Defense Advanced Research Projects Agency DARPA PREEMPT # D18AC00031 and the UCLA AIDS Institute and Charity Treks, and JOL-S was supported by the U.S. National Science Foundation (DEB-1557022), the Strategic Environmental Research and Development Program (SERDP, RCL2635) of the U.S. Department of Defense. Names of specific vendors, manufacturers, or products are included for public health and informational purposes; inclusion does not imply endorsement of the vendors, manufacturers, or products by the US Department of Health and Human Services.

